# Interictal waking and sleep electrophysiological properties of the thalamus in focal epilepsies

**DOI:** 10.1101/2024.03.30.24305106

**Authors:** Tommaso Biagioni, Maria Fratello, Elodie Garnier, Stanislas Lagarde, Romain Carron, Samuel M. Villalon, Isabelle Lambert, Angela Marchi, Julia Makhalova, Agnes Trebuchon, Francesca Bonini, Didier Scavarda, Christian Benar, Fabrice Bartolomei, Francesca Pizzo

**Affiliations:** Department of Clinical and Experimental Medicine, University of Pisa, Pisa, Italy; IRCCS Stella Maris Foundation, Calambrone, Toscana, Italy; INSERM, INS, Inst Neurosci Syst, Aix Marseille Univ, Marseille, France; APHM, Timone Hospital, Epileptology and Cerebral Rhythmology, Marseille, France; APHM, Timone Hospital, Functional, and Stereotactic Neurosurgery, Marseille, France; APHM, Timone Hospital, Sleep Unit, Epileptology and Cerebral Rhythmology, Marseille, France; Aix Marseille Univ, CNRS, CRMBM, Marseille, France; APHM, Department of Pediatric Neurosurgery, Marseille, France

**Author notes:** contributed equally to this work Department of Clinical and Experimental Medicine, University of Pisa, Pisa, Italy IRCCS Stella Maris Foundation, Calambrone, Toscana, Italy Aix Marseille Univ, INSERM, INS, Inst Neurosci Syst, Marseille, France Epileptology and Cerebral Rhythmology, APHM, Timone Hospital, Marseille, France Functional, and Stereotactic Neurosurgery, APHM, Timone Hospital, Marseille, France Sleep Unit, Epileptology and Cerebral Rhythmology, APHM, Timone Hospital, Marseille, France Aix Marseille Univ, CNRS, CRMBM, Marseille, France Pediatric Neurosurgery, APHM, Timone Hospital, Marseille, France **Email:**. **Author Contributions:** Tommaso Biagioni conceptualization, collecting and analyzing data, drafting the manuscript, reviewing and editing the paper for content and clarity, interpreting statistical results, conducting a thorough literature review Maria Fratello collecting and analyzing data, developing or adapting research tools, drafting the manuscript Elodie Garnier analyzing data, drafting the manuscript, conducting statistical analyses, interpreting statistical results Stanislas Lagarde drafting the manuscript, reviewing and editing the paper for content and clarity, reviewing and critiquing the methodology, interpreting statistical results Romain Carron collecting and analyzing data, drafting the manuscript, reviewing and editing the paper for content and clarity Samuel M. Villalon developing or adapting research tools, reviewing and editing the paper for content and clarity Isabelle Lambert drafting the manuscript, reviewing and editing the paper for content and clarity Angela Marchi drafting the manuscript, reviewing and editing the paper for content and clarity Julia Makhalova drafting the manuscript, reviewing and editing the paper for content and clarity Agnes Trebuchon drafting the manuscript, reviewing and editing the paper for content and clarity Francesca Bonini drafting the manuscript, reviewing and editing the paper for content and clarity Didier Scavarda drafting the manuscript, reviewing and editing the paper for content and clarity Christian Benar analyzing data, developing or adapting research tools, interpreting statistical results, drafting the manuscript, reviewing and editing the paper for content and clarity Fabrice Bartolomei* conceptualization, collecting and analyzing data, drafting the manuscript, reviewing and editing the paper for content and clarity, interpreting statistical results Francesca Pizzo* conceptualization, collecting and analyzing data, drafting the manuscript, reviewing and editing the paper for content and clarity, interpreting statistical results, conducting a thorough literature review. **Competing Interest Statement:** The authors declare no competing interests associated with the publication of this paper.

**Keywords:** Intracranial EEG, Thalamic hyperexcitability, Thalamo-cortical functional connectivity, Surgical outcome, Epileptic networks

## Abstract

**Introduction:** Addressing drug-resistant epilepsy poses a significant clinical hurdle, particularly when conventional resective surgery is not feasible. Deep brain stimulation (DBS) emerges as a potential therapeutic avenue for such cases. To shed light on the thalamo-cortical epileptogenic networks and refine prognostic indicators and treatments, we conducted an exhaustive examination of interictal data from thalamic stereoelectroencephalography (SEEG) recordings in a substantial patient cohort (n=121). Our investigation delved into thalamo-cortical connectivity, interictal hyperexcitability biomarkers, and their associations with clinical parameters during both rest and non-rapid eye movement (NREM) sleep states.

**Methodology:** We conducted an observational study retrospectively gathering data from patients who underwent stereotactic-EEG with thalamic implantation as part of pre-surgical evaluation. Interictal recordings during rest and NREM sleep were analyzed to detect spikes and fast ripples automatically. Functional connectivity between the thalamus and other brain regions (involved or non-involved in the epileptogenic network) was examined using linear regression analysis (R2), while statistical evaluations assessed relationships between these measures, patient outcomes, and clinical variables.

**Results:** Elevated rates of thalamic spikes and fast ripples during sleep were linked to unfavorable surgical outcomes (Engel class III/IV) compared to favorable outcomes (Engel class I/II). Thalamo-cortical functional connectivity analysis revealed heightened thalamic strength, particularly in the beta and gamma bands during sleep, among patients with poor surgical outcomes, especially with non-involved networks. Conversely, during rest, lower spike and fast ripple rates were observed in patients with longer epilepsy durations. Furthermore, thalamic strength values during resting state were diminished in patients with prolonged epilepsy durations and older age.

**Discussion:** Our study uncovered heightened thalamic interictal activity and thalamo-cortical functional connectivity during sleep in patients with unfavorable surgical outcomes, within the largest cohort of patients with thalamic recordings documented. These findings inform pre-surgical assessments, aiding in identifying patients unsuitable for surgery and suggesting alternative interventions like DBS. Additionally, diminished thalamic activity and connectivity associated with epilepsy duration and age prompt speculation on the role of the thalamus as a pivotal hub in aging-related processes. Vigilance state-dependent insights from the thalamus underscore the necessity for comprehensive analyses to tailor stimulation protocols, facilitating a personalized therapeutic approach.

## Introduction

Epilepsy, a complex neurological disorder, presents a significant clinical challenge when it proves refractory to antiseizure medications. Deep brain stimulation (DBS) ^1^, associated or not to responsive neurostimulation (RNS) ^2^, is a potential treatment for patients with drug-resistant epilepsy when resective epilepsy surgery is precluded ^3^. To date, DBS of the anterior nucleus of the thalamus is an approved treatment option both in the US (FDA) and Europe (CE-Mark). Despite its increasing adoption, the precise mechanisms governing DBS’s efficacy and identifying optimal patient-specific parameters remain elusive^4^.

The complex relationship between the thalamus and the cortex can be studied using stereoelectroencephalography (SEEG) recording both thalamic and cortical activities during the presurgical evaluation of patients with drug-resistant focal epilepsy ^5–7^. Most of the studies on thalamic SEEG focused on the ictal period, meaning during epileptic seizures ^5,8–10^.

Interestingly, it was observed that the thalamus was involved in seizure in more than 80% of studied cases. The degree of thalamic involvement significantly affected the post-surgical outcome (higher involvement, higher probability of surgical failure) ^9^. Functional connectivity (FC) studies showed a significant involvement of the thalamus in loss of consciousness during seizures (11) and in seizure termination ^10–12^. These data strongly support the thalamus as a pivotal hub in focal epilepsy, underscoring its relevance for comprehending not only the mechanisms of epileptogenicity but also for exploring therapeutic options such as deep brain stimulation (DBS).

Given that DBS often acts in the interictal phase, the study of thalamic electrophysiological characteristics in this period is crucial. However, only a few studies have investigated interictal SEEG thalamic activity and on limited number of patients ^13,14^. Some of these studies tested the response on cortical regions following thalamic stimulation, showing a modification in spike rate and functional connectivity ^15–17^. Regarding cortico-cortical FC in the interictal period, it was evidenced that more extensive FC alterations, implicating the non-involved network and suggesting a more diffuse disease, were associated with poorer post-surgical prognosis ^18^. However, thalamo-cortical interictal FC is still unexplored.

Considering the valuable insights into epileptogenicity, surgical outcomes, and the efficacy of regulating interictal activities derived from the study of thalamic neurophysiology, we postulated that a comprehensive analysis of the intricate relationship between the thalamus and cortex in focal drug-resistant epilepsy, based on a substantial cohort of patients (n=121), could reveal significant correlations with clinical data and potentially identify prognostic factors. We examined thalamic interictal activity and thalamo-cortical interictal functional connectivity during two distinct vigilance states—waking resting state and Non-Rapid Eye Movement (NREM) sleep, which is known to significantly increase interictal epileptic activities ^19^. The deliberate choice of these states provides a nuanced understanding of the dynamic thalamo-cortical interplay across different physiological contexts.

## Materials and Methods

### Patient Selection and clinical data collection

Patients that underwent a stereotactic-EEG study from January 2001 to December 2022 in the Epileptology and Cerebral Rhythmology Unit at the University Hospitals of Marseille, France were selected and included if they had at least one electrode recording from the thalamus. The implantation of the thalamus, most of the time aiming for the pulvinar medialis, is considered when a posterior temporal electrode exploring the *planum temporale* or the Heschl gyrus is needed for the definition of the epileptogenic zone (EZ) and/or for the identification of eloquent cortex. The safety of this procedure has been already discussed in previous studies ^9,20^. Most of the patients in this study were implanted in the pulvinar (88/121) because of the known connections of the medial pulvinar with the limbic structures, the insula, and the lateral temporal cortex ^7^ and because this structure is emerging as a possible site for DBS implantation ^21–24^. Moreover, the pulvinar is easily accessible when it is necessary to explore Heschl’s cortex, in the process of defining language lateralization during the pre-surgical evaluation.

Patient clinical data (date of birth, sex, epilepsy etiology, Epileptogenic Zone (EZ) localization, MRI findings, age at epilepsy onset, and epilepsy duration) and SEEG data (patient’s age at SEEG, localization of the contact exploring the thalamus and eventual bithalamic implantation) were collected. The epilepsy type was defined as: temporal, frontal, occipital, insulo-opercular, parietal and multi-lobar (epilepsies involving more than one cerebral lobe). Finally, data concerning the treatments received during and after the SEEG, such as thermocoagulations, surgery and/or neurostimulation (VNS, DBS or tDCS) were also collected. The surgical outcome has been determined using the Engel classification ^25^, at the last available follow-up, both for patients who underwent surgery and those who received thermocoagulations only.

### Stereotactic-EEG Recordings and interictal recordings

Interictal recordings that were temporally distant from the preceding and the following seizure by at least 2h were selected ^18^. In addition, recordings were chosen at least 2 days after the electrode implantation surgical procedure to limit possible effects of general anesthesia. For each patient were selected:

1. Ten minutes of “rest recording”: time period in which patients stay at rest with eyes closed but awake (recording performed preferable between 07:00 and 12:30 am);
2. Ten minutes of “sleep recording”: N2-N3 sleep stages with minimal number of arousals, possibly during the first or the second sleep cycle. The NREM recordings were selected for the presence of slow waves and spindles in frontal neocortical regions and, notably, in the thalamus.

The stereotactic-EEG exploration was performed in the context of the presurgical evaluation of epilepsy using intracerebral multiple-contact electrodes (Dixi Medical or Alcis), consisting of 10–18 contacts with length 2 mm, diameter 0.8 mm, spaced by 1.5mm ^9^. Signals were recorded on a 128- or 256-channel Natus system depending on the year of recording. They were sampled at 256, 512, 1024, or 2048 Hz depending on the year of recording, and recorded on a hard disk (16 bits/sample) using no digital filter. Two hardware filters were present in the acquisition procedure: a high-pass filter (cut-off frequency = 0.16 Hz at -3 dB), and an anti-aliasing low-pass filter (cut-off frequency = 97 Hz for 256 Hz sampling rate, 170 Hz for 512 Hz sampling rate, 340 Hz for 1024Hz sampling rate, or 680 Hz for 2048 Hz sampling rate). Artifacts and bad channels were excluded from the analysis.

### BIDS Anywave

The anonymized clinical data of the patients, their imaging and SEEG records were organized according to the BIDS (Brain Imaging Data Structure) standard (24), using BIDS Manager ^26^, a software that allows to explore and dynamically organize the data according to the BIDS standard whilst tracking the subject data readiness and integrity.

All signal analysis was computed using the Anywave software ^27^ (open-source at http://meg.univ-amu.fr/wiki/AnyWave).

### Semi-automatic Interictal Activities Detection

To detect interictal paroxysmal activities during rest and sleep, the Delphos detector was used, looking for spikes (all recordings) and fast ripple high frequency oscillations (250-500 Hz) (for recordings sampled at 2048 Hz) on all channels without artifacts. The analysis was conducted on a bipolar montage. The Delphos detector ^28^ is based on the ZH0 method, which aims at flattening the frequency spectrum to enhance the fast oscillations while preserving an optimal signal-to-noise ratio at each frequency. In this study, the ripple frequencies (150-250 Hz) were not studied, in order to avoid the possible interferences of physiological activities in that frequency band ^29^.

Because some patients had multiple bipolar channels recording the thalamus, we computed at the patients level the maximum thalamic Spike rate and HFOs rate, during rest and sleep (called “Spike Max” and “HFO Max”). Furthermore, as these rates were highly variable from individual to individual, all values were normalized by the highest Spike or HFO rate (across at SEEG channels) recorded in the individual patient and therefore the values are between 0 and 1.

### Functional Connectivity Analysis: Liner regression method (R^2^)

To investigate the functional connectivity between the thalamus and the other brain regions during the interictal period (rest and sleep), we computed on bipolar montage the linear correlation coefficient R^2^. Despite acknowledging the non-linear properties inherent in neuronal network dynamics, especially in epilepsy ^30^, the study opted for a linear regression method due to its computational efficiency compared to non-linear regression methods. The ongoing debate about the superiority of non-linear methods, which, in certain situations, demonstrated comparable performance to linear methods ^31^, factored into this decision. For two time series x (t) and y (t), Pearson correlation coefficient is defined in the time domain as:

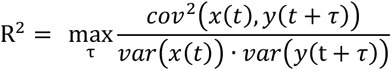

where var, cov, and τ denote respectively variance, covariance, and time shift between the two time series ^32^. Correlation values are contained between 0 (y is uncorrelated with x) and 1 (x and y are fully linearly dependent). The estimation of this parameter is performed on a temporal window of fixed duration and sliding in time in order to follow the temporal evolution of the linear statistical relationship between both signals ^31^. The method investigates also the effect of a shifting temporally the two signals and retained the maximal R^2^ value across all explored time-shift. In the present study the following parameters for R^2^ analysis were used: time window 4 sec, maximum time-shift 0.1 sec, steps 1 sec. The functional connectivity analysis was conducted on a grey matter bipolar montage, excluding white matter channels. When constructing the connectivity matrix, the values that referred to the connection of intra-thalamic channels were removed to highlight the connections between the thalamus and the cortical areas. For each thalamic bipolar channel, we first average R^2^ values trough the time windows. Then the node strength was computed, by summing the R^2^ values between the thalamic bipolar channel and all the other non-thalamic bipolar channels and normalizing by the number of non-thalamic bipolar channels. In order to obtain one single thalamic strength value for patient’s recording (one for rest and one for sleep), the mean between the node strength of the available thalamic channels was calculated, which is referred to as “thalamic strength”. The analysis was firstly conducted on all thalamic channels and later considering pulvinar channels only, obtaining similar results. The analysis was realized on broadband signal, but also on raw signals filtered in classically defined EEG sub-bands: delta (0.5-3.4 Hz), theta (3.4-7.4 Hz), alpha (7.4-12.4 Hz), beta (12.4-24 Hz) and gamma (24-80 Hz) ^18^.

Analyses of the functional connectivity results was also investigated according to the epileptogenicity of the structure. Epileptogenicity was determined using the Epileptogenic Index ^33^ that is a semi-automatic method to quantify the ictal discharges ^34^. Briefly, EI estimates the propensity of a brain region to generate fast discharge early during the seizure course. Channels were considered in the Epileptogenic zone (EZ) if the EI > 0.4, in the Propagation zone (PZ) if EI included between 0.1 and 0.4 and in the NIZ if EI <0.1.

### Statistical Analyses

To evaluate the relationships between the paroxysmal interictal activities recorded in the thalamus (spikes and fast ripples) and the thalamic functional connectivity measure (thalamic strength) and patient’s surgical outcome, primary objectives of this study, the Spearman rank correlation was used. Because of the low number of patients being Engel IV after surgery (11 patients, but 9 who received thermocoagulations), Anova with Wilcoxon test was performed (after finding a non-normal distribution of the variables) considering only two possible measures of outcome, differenced by the persistence of invalidating seizures (Engel III and IV *vs* Engel II and I). Additionally, the association of interictal activities and thalamic strength with the clinical variables was studied. Similarly, for continuous variables such as “duration of epilepsy” and “age at SEEG” the Spearman rank correlation was performed; for categorial variables as “etiology”, “epilepsy type”, and “lateralization”, Anova with Wilcoxon test was used. The significance for multiple comparisons was corrected using the False Discovery Rate method (FDR). Finally, the correlation between Engel class and duration of epilepsy was calculated, and, in the same way, the correlation between Engel class and age at the moment of SEEG, using the Spearman rank correlation test. A multivariate analysis to evaluate the correlation between age, duration of epilepsy and surgical outcome was performed. The statistical analyses were performed using the software R.

### Data availability

Data are available upon reasonable request.

Standard Protocol Approvals, Registrations, and Patient Consents

Written patient consent was obtained both for the implantation of the electrodes for SEEG and for the subsequent RF-TC procedures; the study has received approval from the review board (WFE8Q9).

## Results

### Hyperexcitability of the thalamus

To investigate the level of thalamic hyperexcitability during interictal periods, we examined the rates of interictal paroxysmal biomarkers. Specifically, we focused on two key biomarkers: spikes (as illustrated in Figure 1) and high-frequency oscillations (HFOs) ^35^, with a specific emphasis on fast ripples, defined as oscillations occurring within the frequency range of 250 to 500 Hz ^36^. An automatic detector was employed to systematically analyze and quantify these biomarkers ^28^. The distribution of Spike and HFOs for the patients is reported in Supplementary Figure 1.

**Figure 1.**
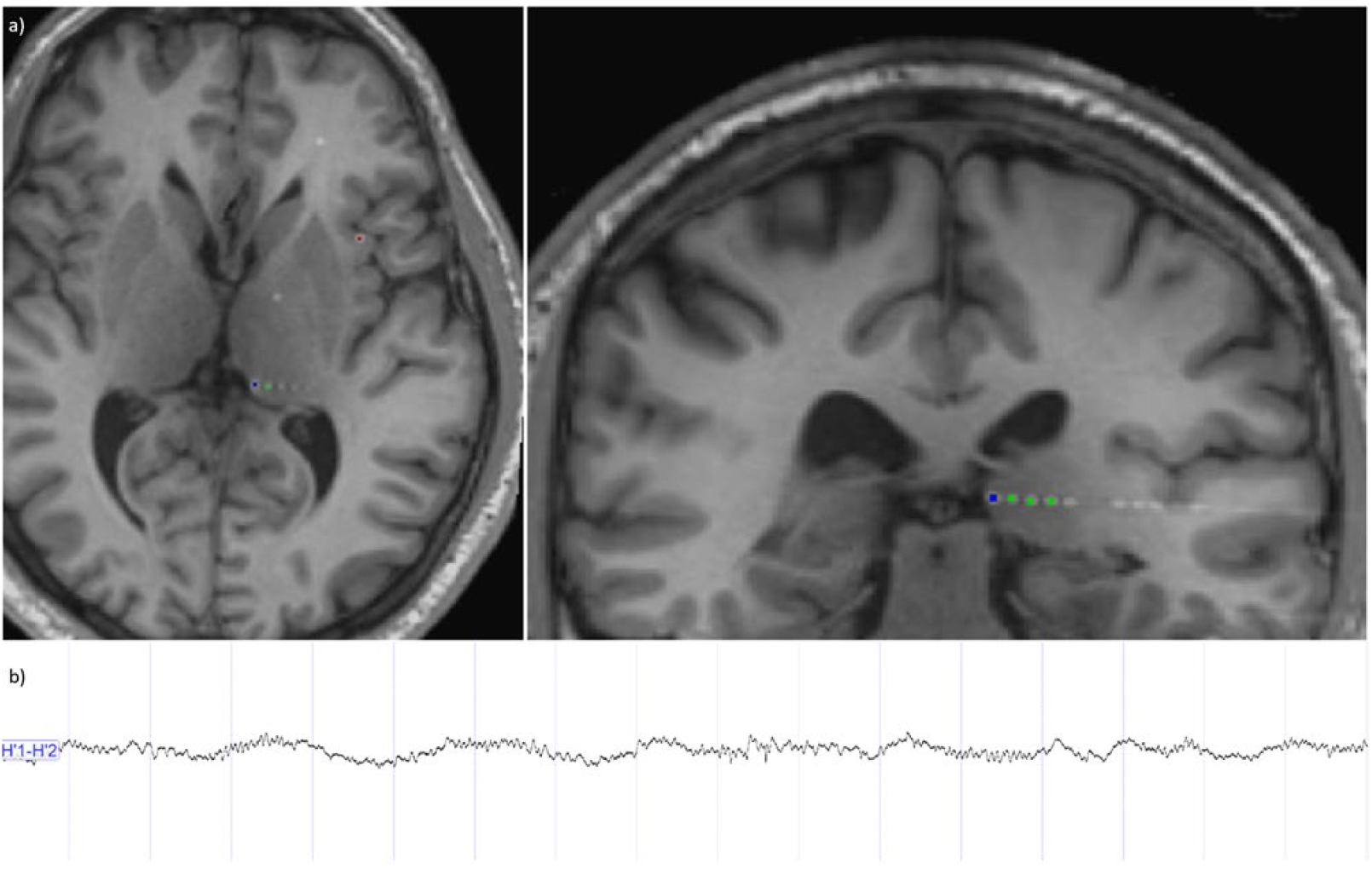
Thalamic implantation and signal recordings. a) MRI/CT co-registration showing a SEEG electrode whose distal contacts are implanted at the level of the left pulvinar. b) A SEEG signal was recorded at the level of the two distal contacts of the electrode implanted in the pulvinar, in a sleep recording, recognizable for slow wave and spindle activity. The presence of interictal epileptiform activity, a spike, is observed (arrow).

One-hundred-twenty-one patients (59 females) were analyzed in this study, the clinical features are detailed in Table 1.

**Table 1.**
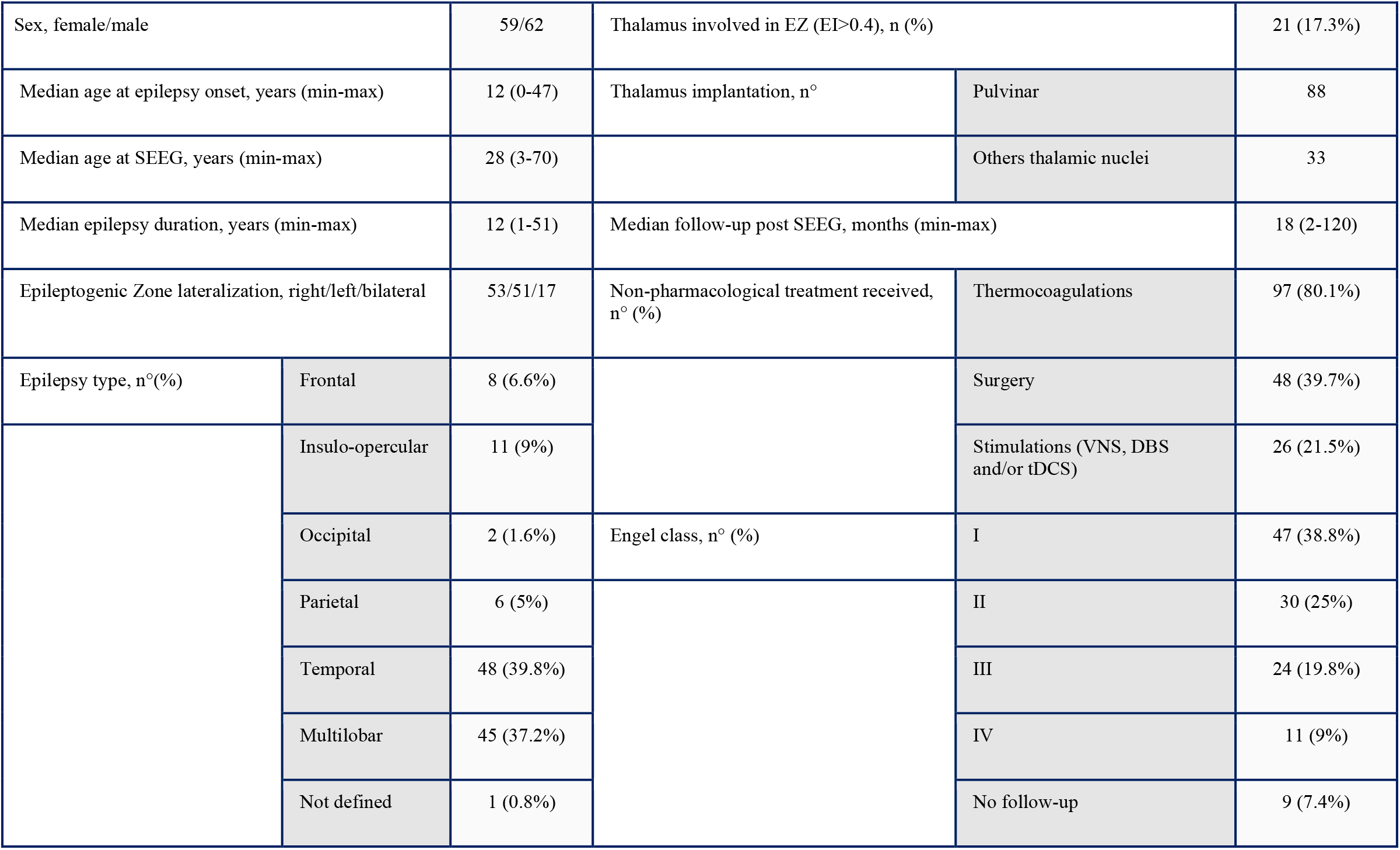
Clinical characteristics of the cohort

The investigation aimed to establish a correlation between hyperexcitability biomarker rates and surgical outcomes, as determined by the Engel score ^25^. We found a higher thalamic spike rate during sleep in patients with bad surgical outcome (Engel class III/IV) compared to those with good outcome (Engel class I/II) (Wilcoxon test, p<0.01, Fig 2a). Additionally, the rate of thalamic high-frequency oscillations (HFOs), specifically fast ripples (250-500 Hz), during sleep was also higher in patients with bad surgical outcome (compared to those with good outcome, Wilcoxon test, p<0.05, Fig 2b). We also observed a slight increasing trend in the HFOs rate with the severity of Engel classes (Fig 2c). Importantly, these associations were observed only during sleep recordings and not during rest. The findings suggest that thalamic hyperexcitability during sleep, as measured by interictal paroxysmal biomarkers, may serve as a potential new marker for predicting epilepsy outcomes after surgery.

**Figure 2.**
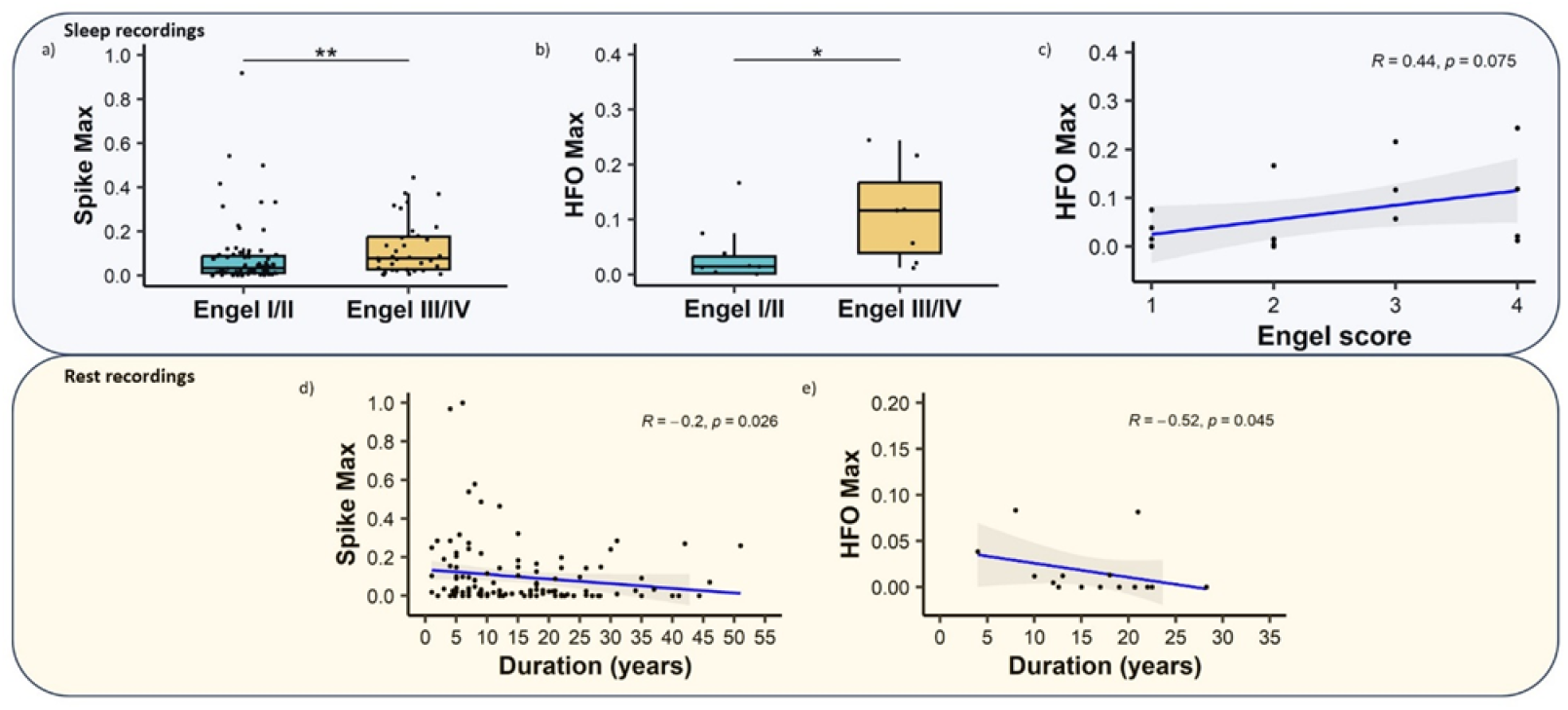
Thalamic interictal biomarkers associated with hyperexcitability and clinical characteristics: Association between surgical outcome and spikes (a) and fast ripples (b) in sleep recordings. Fast ripples rate according to Engel score (c). Association between epilepsy duration and spikes (d) and fast ripples (e) in rest recordings.

During rest recordings, we observed a reduction of interictal paroxysmal activities, both in terms of spike and fast ripple (FR) rates (p<0.05 in both cases, as depicted in Fig 2d and 2e), in patients with loger duration of epilepsy. This suggests a potential relationship between thalamic hyperexcitability and the temporal evolution of epilepsy. We also checked the possible correlation between Engel class and duration of epilepsy, and, in the same way, the correlation between Engel class and age at the moment of SEEG, without finding any significancy.

However, conversely, no differences were found in spike and high-frequency oscillations (HFOs) rates concerning various factors such as age at SEEG, epilepsy type, lateralization, etiology and thalamic involvement in the Epileptogenic Zone (defined by Epileptogenic Index values > 0.4 in the thalamic channels)^33^.

### Thalamo-cortical functional connectivity

To examine the functional relationships between the thalamus and the cortical regions, particularly considering their roles in the ictal epileptogenic network (defined within epileptogenic zone, propagation zone, or non-involved zone) ^37^, a functional connectivity analysis was conducted using linear correlation coefficient (R^2^). The study posited that if thalamic functional connectivity (FC) is linked to surgical outcomes, modulating the network involving this central “hub” through interventions such as DBS might alter the network organization and potentially modify the clinical course.

Analysis of thalamic strength during sleep recordings showed higher values for patients with bad postsurgical outcome (Engel class III/IV) compared to those with a good-one (Engel class I/II). Notably, these differences were observed in the beta and gamma bands (p<0.001 and p<0.5, respectively; depicted in Fig 3a and 3b), but not in broadband and other sub-bands (delta, theta, alpha). Furthermore, in the beta band, a discernible increasing trend in thalamic strength values with the severity of Engel classes was noted (p<0.01; Fig 3c).

**Figure 3.**
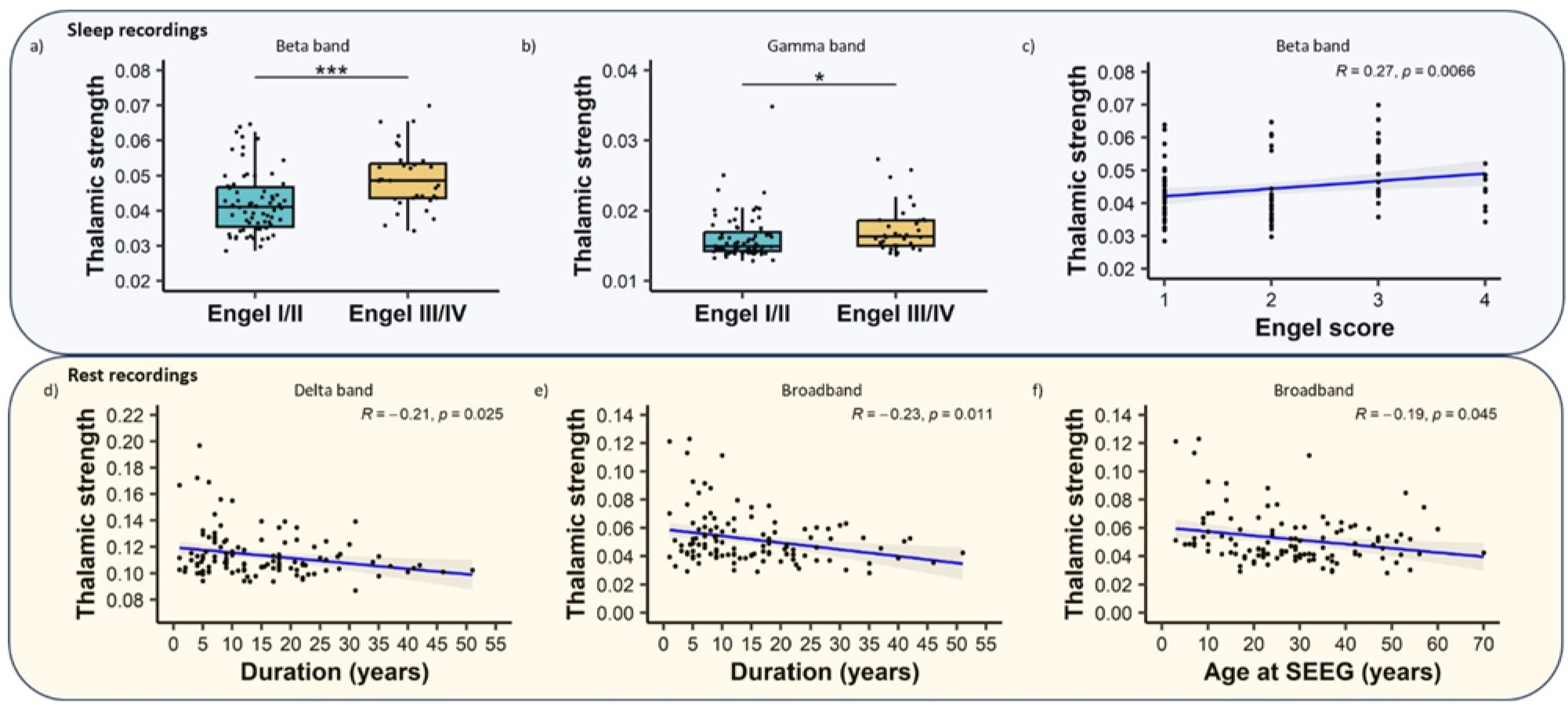
Thalamo-cortical interictal functional connectivity and clinical characteristics. Association between surgical outcome and functional connectivity in beta (a) and gamma (b) band in sleep recordings. Functional connectivity in beta band varies according to Engel score (c). Association between epilepsy duration and thalamo-cortical functional connectivity in delta (d) and broad (e) band in rest recordings. Association between age at SEEG and thalamo-cortical functional connectivity in broadband (f) in rest recordings.

It has been further explored if thalamic functional connectivity (FC) was different according to the sub-networks (EZ, PZ, NI). We found higher connectivity with the non-involved network (NI) in sleep recordings in the beta and gamma bands (p<0.01 and p<0.01) in patients with higher Engel class. Therefore, connections within the non-involved networks emerged as more influential on the thalamo-cortical correlation with surgical outcomes. Importantly, none of these correlations with surgical outcome were identified during rest recordings.

In addition, thalamic strength values reduced with the duration of epilepsy during rest recordings, in both delta band and broadband (p<0.05; Fig 3d and 3e). A similar thalamic strength values reduction was also identified for older patients (higher age at SEEG) during rest recordings in broadband (p<0.05; Fig 3f). No differences for thalamic strength were found regarding to etiology, epilepsy type, and lateralization.

Similarly to sleep recordings, when examining thalamic FC in rest recordings according to the sub-networks (EZ, PZ, NI), we found a reduction following the duration of epilepsy (p<0.01) and age at SEEG (p<0.05) were specifically evident with the non-involved zone (NIZ) and not with the involved zone, obtained merging the propagation zone (PZ) and the epileptogenic zone (EZ).

## Discussion

In this study, findings from an extensive cohort of 121 patients with thalamic SEEG recordings are presented, marking the most comprehensive analysis to date. This exploration of interictal SEEG thalamic recordings not only sheds light on their clinical relevance in predicting post-surgical seizure outcomes, but also contributes to the broader understanding of subcortical-cortical interactions in focal epilepsies.

A notable aspect of the study lies in the differentiation of thalamic properties based on the patient’s vigilance state—awake or during NREM sleep. These states provide distinct information offering insights into surgical outcome (sleep) and into the disease course and evolution (awake rest). This suggests that the thalamus harbors critical information for both the progression of the disease and the ability to predict surgical outcomes. Thus, these observations reinforce the pivotal role of the thalamus in focal epilepsy, supporting the modulation by DBS approaches for influencing epileptogenic network dynamics.

Analysis of functional connectivity revealed informative patterns, particularly in thalamic connections with cortical areas outside the epileptogenic network. This complexity underscores the intricate organization of focal epilepsies and potential explanations for varied neurocognitive and neuropsychological comorbidities. Building on prior work investigating cortical functional connectivity in the interictal state ^18^, where the Non-Involved network (NI) played a crucial role in predicting surgical outcomes, current findings reaffirm the broad and nuanced nature of what is conventionally labeled as “focal” epilepsy.

### Thalamic hyperexcitability biomarkers and functional connectivity during sleep are linked to the surgical outcome

This study revealed a compelling association between hyperexcitability biomarkers (higher rate of spikes and FR) in the thalamus during sleep recordings and unfavorable surgical outcomes. Few previous studies reported the presence of hyperexcitability biomarkers in the thalamus, mainly spikes ^6^ and in small cohort of patients ^13^. In this last study ^13^, the presence of thalamic spikes was associated with a worst DBS outcome. These results are consistent with the notion that the extension of the spike network could be a prognosis factor in patients operated after SEEG ^38^. However, it is probably an indices of the extension of the epileptogenicity, having in mind that surgical outcome after SEEG is more related to ictal than interictal biomarkers ^39^.

Concerning FR in thalamus, a scarcity of data in human studies prompted this investigation ^14^. Although FR detection was limited to a small number of patients (rest n=15, sleep n=17) due to sampling rate constraints, this study provides valuable insights into their potential significance as outcome predictor, underscoring the need for further exploration.

This investigation, employing the R^2^ linear correlation method to explore functional connectivity, reveals also an association of higher values of functional connectivity in the thalamus, particularly during sleep recordings in the beta- and gamma-bands, and worse surgical outcomes. This observation aligns seamlessly with neuroimaging studies, where patients experiencing persistent seizures post temporal lobe epilepsy surgery exhibited notable white matter abnormalities in thalamo-temporal tracts ^40^, disrupted right thalamo-hippocampus resting state functional connectivity ^41^, and a specific increase in nodal hubness involving both ipsilateral and contralateral thalami ^42^. Our results add a new dimension to this understanding by demonstrating that the thalamo-cortical functional connectivity on SEEG, particularly with the NIZ, plays a pivotal role in predicting surgical outcomes. This observation is in line with previous works showing relationship between altered functional connectivity extent and surgical outcome ^43^.

Notably, hyperexcitability biomarkers in the mesial temporal region, are profoundly shaped by vigilance states ^44^ prompting their exploration within the thalamus. Recent studies suggest NREM sleep as the best vigilance state to identify the epileptogenic zone network ^45,46^ establishing it as a critical period for prognostic insights. The impact of spikes during sleep ^47^ extends beyond the nocturnal realm, influencing neurological performance during wakefulness ^48^. The findings of this study, aligned with previous research on neocortical regions, establish correlations between hyperexcitability biomarkers and surgical prognosis in the thalamus, specifically during sleep.

In summary, the findings of this study underscore the critical role of thalamic hyperexcitability biomarkers and thalamo-cortical functional connectivity during sleep in predicting surgical outcomes in focal drug-resistant epilepsy. This insight, derived from neurophysiological studies, enriches the understanding of the complex dynamics governing the efficacy of surgical interventions in epilepsy.

### Thalamic hyperexcitability biomarkers and FC decrease with epilepsy duration and age during rest recordings

During resting-state recordings, a distinctive correlation between the duration of epilepsy and changes in thalamic interictal paroxysmal activity was found, suggesting a reduction in functional connectivity over time. Specifically, a progressive reduction was observed in hyperexcitability biomarkers measured in the thalamus, potentially linked to a concurrent reduction in functional connectivity with increasing epilepsy duration.

An inverse relationship between duration of epilepsy and EEG functional connectivity has been demonstrated in other studies on intracranial ^49^ and surface neurophysiological recordings ^50^. Changes in SEEG network topology in relationship to epilepsy duration has been also demonstrated in the past ^51^. Moreover, several MRI studies have shown variations in white matter volume ^52^ and abnormal fractional anisotropy (between the homolateral insula and the superior temporal pole) ^53^ associated with the duration of epilepsy. Longer durations of epilepsy have also been associated with lower subcortical volume, including thalamus, and cortical thickness ^54^. This research confirms possible effects of epilepsy duration on functional connectivity involving the thalamus.

In parallel, age-related differences in thalamo-cortical connectivity were found, a topic less explored in the context of epilepsy. The existing literature suggests that brain’s functional and structural connectivity patterns decrease with older age in normal subjects ^55^ and in a more pronounced way in patients with Alzheimer disease, in particular in the default mode network ^56^. Focusing on the thalamus, age-related differences in thalamo-cortical connectivity may contribute to age-related changes in attention, working memory and episodic memory processes ^57^.

The SEEG recordings also revealed a lower thalamo-cortical functional connectivity in older patients, independent of epilepsy duration. This reduction during resting state recordings hints at a potential parallel with the well-documented fMRI-default network modification with age.

### Methodological aspects and considerations

Strength of this study is the semi-automation of the signal analysis process. Indeed, after rest and sleep recordings selection according to the criteria described in the methods section, and exclusion of bad-channels and artifacts, the analysis of interictal activity and functional connectivity were performed automatically on all patients and with an open source software (http://meg.univ-amu.fr/wiki/AnyWave) promoting reproducibility.

Concerning the surgical outcome, one of the major limitations of our study is certainly that we considered patients who underwent conventional surgery and patients who underwent thermocoagulations during SEEG in the same way. This choice was tied to the conceptual similarity between surgery and thermocoagulation regarding the potential role of the thalamus as subcortical hub. However, to confirm the results obtained, we repeated the analysis only on patients who had received thermocoagulations during the SEEG and not subsequent surgery, finding results that completely overlapped with those described. On the other hand, it was not possible to repeat the analysis considering patients who had received only conventional surgery, because in our case series there are only 2 patients (2 pts Engel III, 0 pts Engel IV) who continued to have disabling seizures after surgery, preventing a statistical comparison with seizure-free patients after surgery (32 pts Engel I, 11 pts Engel II).

Another possible bias in this study is related to the anti-seizure medications (ASMs) that the patients were taking during their hospitalisation to record the SEEG. Usually, ASMs were withdrawn during the SEEG recording to facilitate seizures. Therefore, considering that we selected two SEEG recordings (rest and N-REM) for each patient, it is possible that the recordings for each patient were influenced by different ASMs or concentrations of these drugs.

### Conclusions

This study reaffirms the importance of investigating the thalamus within the epileptogenic networks in focal epilepsies. The thalamus exhibits heightened interictal activity (both Spikes and HFO) and functional connectivity during sleep in patients with poor surgical outcomes compared to those with favorable outcomes after surgery and/or thermocoagulations. These findings present a valuable tool for clinicians in pre-surgical assessments, aiding in the identification of patients unsuitable for surgery and potentially guiding them towards alternative clinical interventions, such as Deep Brain Stimulation (DBS).

Additionally, this study highlights a reduction in interictal activity and functional connectivity within the thalamus associated with epilepsy duration and age in rest recordings. This prompts speculation about the implications of persistent epilepsy on brain networks and the possible role of the thalamus as a central hub of connections in the aging process.

In conclusion, the thalamus provides information differently depending on the vigilance state, with more epilepsy-related insights unearthed during slow-wave sleep. Conversely, during the resting state, thalamic neurophysiology appears more linked to the “default mode network”, the aging of patients and epilepsy evolution.

Further analyses, incorporating both resting and sleep SEEG recordings, are essential to uncover specific connections between the thalamus and distinct brain areas. This detailed exploration will contribute to identifying targets for tailoring stimulation protocols, offering a more personalized approach to each patient.

## Data Availability

All data produced in the present study are available upon reasonable request to the authors

## Acknowledgments

SL is supported by grants from LFCE (Ligue Française Contre l’Epilepsie, French chapter of the ILAE) and PHOCEO.

## References

1. Ryvlin P, Rheims S, Hirsch LJ, Sokolov A, Jehi L. Neuromodulation in epilepsy: state-of-the-art approved therapies. Lancet Neurol. 2021;20(12):1038–1047. doi:10.1016/S1474-4422(21)00300-8

2. Nair DR, Morrell MJ, Skarpaas TL, et al. Nine-year prospective efficacy and safety of brain-responsive neurostimulation for focal epilepsy. Neurology. 2020;95(9):E1244–E1256. doi:10.1212/WNL.0000000000010154

3. Touma L, Dansereau B, Chan AY, et al. Neurostimulation in people with drug-resistant epilepsy: Systematic review and meta-analysis from the ILAE Surgical Therapies Commission. Epilepsia. 2022;63(6):1314–1329. doi:10.1111/epi.17243

4. Piper RJ, Richardson RM, Worrell G, et al. Towards network-guided neuromodulation for epilepsy. Brain. 2022;145(10):3347–3362. doi:10.1093/brain/awac234

5. Guye M, Régis J, Tamura M, et al. The role of corticothalamic coupling in human temporal lobe epilepsy. Brain. 2006;129(7):1917–1928. doi:10.1093/brain/awl151

6. Wu TQ, Kaboodvand N, McGinn RJ, et al. Multisite thalamic recordings to characterize seizure propagation in the human brain. Brain. 2023;146(7):2792–2802. doi:10.1093/brain/awad121

7. Rosenberg DS, Mauguière F, Catenoix H, Faillenot I, Magnin M. Reciprocal thalamocortical connectivity of the medial pulvinar: A depth stimulation and evoked potential study in human brain. Cereb Cortex. 2009;19(6):1462–1473. doi:10.1093/cercor/bhn185

8. Romeo A, Issa Roach AT, Toth E, et al. Early ictal recruitment of midline thalamus in mesial temporal lobe epilepsy. Ann Clin Transl Neurol. Published online 2019. doi:10.1002/acn3.50835

9. Pizzo F, Roehri N, Giusiano B, et al. The Ictal Signature of Thalamus and Basal Ganglia in Focal Epilepsy: A SEEG Study. Vol 96.; 2021. doi:10.1212/WNL.0000000000011003

10. Ilyas A, Toth E, Chaitanya G, Riley K, Pati S. Clinical Neurophysiology Ictal high-frequency activity in limbic thalamic nuclei varies with electrographic seizure-onset patterns in temporal lobe epilepsy. Clin Neurophysiol. 2022;137:183–192. doi:10.1016/j.clinph.2022.01.134

11. Evangelista E, Bénar C, Bonini F, et al. Does the thalamo-cortical synchrony play a role in seizure termination? Front Neurol. 2015;6(SEP). doi:10.3389/fneur.2015.00192

12. Soulier H, Pizzo F, Jegou A, et al. The anterior and pulvinar thalamic nuclei interactions in mesial temporal lobe seizure networks. Clin Neurophysiol. 2023;150:176–183. doi:10.1016/j.clinph.2023.03.016

13. Sweeney-Reed CM, Lee H, Rampp S, et al. Thalamic interictal epileptiform discharges in deep brain stimulated epilepsy patients. J Neurol. 2016;263(10):2120–2126. doi:10.1007/s00415-016-8246-5

14. Rektor I, Doležalová I, Chrastina J, et al. High-Frequency Oscillations in the Human Anterior Nucleus of the Thalamus. Brain Stimul. 2016;9(4):629–631. doi:10.1016/j.brs.2016.04.010

15. Velasco M, Velasco F, Velasco AL, Luján M, del Mercado JV. Epileptiform EEG Activities of the Centromedian Thalamic Nuclei in Patients with Intractable Partial Motor, Complex Partial, and Generalized Seizures. Epilepsia. 1989;30(3):295–306. doi:10.1111/j.1528-1157.1989.tb05301.x

16. Labar D, Dakov P, Kobylarz E, Nikolov B, Schwartz TH, Fisher S. Effects of responsive electrical brain stimulation on intracranial electroencephalogram spikes. Neuromodulation. 2013;16(4):355–361. doi:10.1111/ner.12039

17. Yu T, Wang X, Li Y, et al. High-frequency stimulation of anterior nucleus of thalamus desynchronizes epileptic network in humans. Published online 2018:2631–2643. doi:10.1093/brain/awy187

18. Lagarde S, Roehri N, Lambert I, et al. Interictal stereotactic-EEG functional connectivity in refractory focal epilepsies. 2018;(August):1–15. doi:10.1093/brain/awy214

19. Sammaritano M, Gigli GL, Gotman J. Interictal spiking during wakefulness and sleep and the localization of foci in temporal lobe epilepsy. Neurology. 1991;41(2 (Pt 1)):290–297. doi:10.1212/wnl.41.2_part_1.290

20. Chaitanya G, Romeo AK, Ilyas A, et al. Robot-assisted stereoelectroencephalography exploration of the limbic thalamus in human focal epilepsy: Implantation technique and complications in the first 24 patients. Neurosurg Focus. 2020;48(4):1–11. doi:10.3171/2020.1.FOCUS19887

21. Filipescu C, Lagarde S, Lambert I, et al. The effect of medial pulvinar stimulation on temporal lobe seizures. Epilepsia. 2019;60(4)(Avril):e25–e30. doi:10.1111/epi.14677

22. Burdette D, Mirro EA, Lawrence M, Patra SE. Brain-responsive corticothalamic stimulation in the pulvinar nucleus for the treatment of regional neocortical epilepsy: A case series. Epilepsia Open. 2021;6(3):611–617. doi:10.1002/epi4.12524

23. Vakilna YS, Chaitanya G, Hafeez MU, et al. Pulvinar neuromodulation for seizure monitoring and network modulation in temporal plus epilepsy. Ann Clin Transl Neurol. Published online May 2023:1–6. doi:10.1002/acn3.51815

24. Deutschová B, Pizzo F, Giusiano B, et al. Ictal connectivity changes induced by pulvinar stimulation correlate with improvement of awareness. Brain Stimul. 2021;14(2):344–346. doi:10.1016/j.brs.2021.01.021

25. Engel J. Update on surgical treatment of the epilepsies. Clin Exp Neurol. 1992;29(August):32–48.

26. Roehri N, Medina Villalon S, Jegou A, et al. Transfer, Collection and Organisation of Electrophysiological and Imaging Data for Multicentre Studies. Neuroinformatics. 2021;19(4):639–647. doi:10.1007/s12021-020-09503-6

27. Colombet B, Woodman M, Badier JM, Bénar CG. AnyWave: A cross-platform and modular software for visualizing and processing electrophysiological signals. J Neurosci Methods. 2015;242:118–126. doi:10.1016/j.jneumeth.2015.01.017

28. Roehri N, Pizzo F, Bartolomei F, Wendling F, Bénar CG. What are the assets and weaknesses of HFO detectors? A benchmark framework based on realistic simulations. PLoS One. 2017;12(4). doi:10.1371/journal.pone.0174702

29. Frauscher B, von Ellenrieder N, Zelmann R, et al. High-frequency oscillations in the normal human brain. Ann Neurol. Published online 2018. doi:10.1002/ana.25304

30. Jirsa VK, Stacey WC, Quilichini PP, Ivanov AI, Bernard C. On the nature of seizure dynamics. Brain. 2014;137(Pt 8):2210–2230. doi:10.1093/brain/awu133

31. Ansari-Asl K, Wendling F, Bellanger JJ, Senhadji L. Comparison of two estimators of time-frequency interdependencies between nonstationary signals: application to epileptic EEG. Conf Proc . Annu Int Conf IEEE Eng Med Biol Soc IEEE Eng Med Biol Soc Annu Conf. 2004;2006:263–266. doi:10.1109/IEMBS.2004.1403142

32. Ansari-Asl K, Senhadji L, Bellanger JJ, Wendling F. Quantitative evaluation of linear and nonlinear methods characterizing interdependencies between brain signals. Phys Rev E Stat Nonlin Soft Matter Phys. 2006;74(3 Pt 1):31916. doi:10.1103/PhysRevE.74.031916

33. Bartolomei F, Chauvel P, Wendling F. Epileptogenicity of brain structures in human temporal lobe epilepsy: A quantified study from intracerebral EEG. Brain. 2008;131(7):1818–1830. doi:10.1093/brain/awn111

34. Medina Villalon S, Paz R, Roehri N, et al. EpiTools, A software suite for presurgical brain mapping in epilepsy: Intracerebral EEG. J Neurosci Methods. 2018;303:7–15. doi:10.1016/j.jneumeth.2018.03.018

35. Roehri N, Pizzo F, Lagarde S, et al. High-frequency oscillations are not better biomarkers of epileptogenic tissues than spikes. Ann Neurol. 2018;83(1):84–97. doi:10.1002/ana.25124

36. Nevalainen P, Von Ellenrieder N, Klimeš P, Dubeau F, Frauscher B, Gotman J. Association of Fast Ripples on Intracranial EEG and Outcomes after Epilepsy Surgery. Vol 95.; 2020. doi:10.1212/WNL.0000000000010468

37. Bartolomei F, Lagarde S, Wendling F, et al. Defining epileptogenic networks: Contribution of SEEG and signal analysis. Epilepsia. 2017;58(7):1131–1147. doi:10.1111/epi.13791

38. Luan C, Miller J, Sollars C, Peng J, Singh J. Stereo-Electroencephalography-Recorded Interictal Epileptiform Discharges: Activation Pattern and Its Relationship With Surgical Outcome. Cureus. 2023;15(7):4–8. doi:10.7759/cureus.41337

39. Makhalova J, Madec T, Medina Villalon S, et al. The role of quantitative markers in surgical prognostication after stereoelectroencephalography. Ann Clin Transl Neurol. 2023;10(11):2114–2126. doi:10.1002/acn3.51900

40. Keller SS, Richardson MP, O’Muircheartaigh J, Schoene-Bake JC, Elger C, Weber B. Morphometric MRI alterations and postoperative seizure control in refractory temporal lobe epilepsy. Hum Brain Mapp. 2015;36(5):1637–1647. doi:10.1002/hbm.22722

41. Morgan VL, Sonmezturk HH, Gore JC, Abou-Khalil B. Lateralization of temporal lobe epilepsy using resting functional magnetic resonance imaging connectivity of hippocampal networks. Epilepsia. 2012;53(9):1628–1635. doi:10.1111/j.1528-1167.2012.03590.x

42. He X, Doucet GE, Pustina D, Sperling MR, Sharan AD, Tracy JI. Presurgical thalamic hubness predicts surgical outcome in temporal lobe epilepsy. Neurology. 2017;88(24):2285–2293. doi:10.1212/WNL.0000000000004035

43. Lagarde S, Benar CG, Wendling F, et al. Interictal Functional Connectivity in Focal Refractory Epilepsies Investigated by Intracranial EEG. Brain Connect. 2022;00(00). doi:10.1089/brain.2021.0190

44. Lambert I, Roehri N, Giusiano B, et al. Brain regions and epileptogenicity influence epileptic interictal spike production and propagation during NREM sleep in comparison with wakefulness. Epilepsia. 2017;(October):1–9. doi:10.1111/epi.13958

45. Frauscher B, Von Ellenrieder N, Ferrari-Marinho T, Avoli M, Dubeau F, Gotman J. Facilitation of epileptic activity during sleep is mediated by high amplitude slow waves. Brain. 2015;138(6):1629–1641. doi:10.1093/brain/awv073

46. Klimes P, Cimbalnik J, Brazdil M, et al. NREM sleep is the state of vigilance that best identifies the epileptogenic zone in the interictal electroencephalogram. Epilepsia. 2019;60(12):2404–2415. doi:10.1111/epi.16377

47. Nobili L, Frauscher B, Eriksson S, et al. Sleep and epilepsy: A snapshot of knowledge and future research lines. J Sleep Res. 2022;(April):1–11. doi:10.1111/jsr.13622

48. Lambert I, Tramoni-negre E, Lagarde S, et al. Accelerated long-term forgetting in focal epilepsy : Do interictal spikes during sleep matter ? Epilepsia. 2021;(January):1–7. doi:10.1111/epi.16823

49. van Dellen E, Douw L, Baayen JC, et al. Long-Term Effects of Temporal Lobe Epilepsy on Local Neural Networks: A Graph Theoretical Analysis of Corticography Recordings. PLoS One. 2009;4(11). doi:10.1371/journal.pone.0008081

50. Zhang J, Npeng, Feng J, et al. Resting-State MEG Source Space Network Metrics Associated with the Duration of Temporal Lobe Epilepsy. Brain Topogr. 2021;6:731–744.

51. Bartolomei F, Wendling F, Régis J, Gavaret M, Guye M, Chauvel P. Pre-ictal synchronicity in limbic networks of mesial temporal lobe epilepsy. Epilepsy Res. 2004;61(1-3):89–104. doi:10.1016/j.eplepsyres.2004.06.006

52. Xu S wen, Xi J hui, Lin C, et al. Cognitive decline and white matter changes in mesial temporal lobe epilepsy. Med (United States). 2018;97(33). doi:10.1097/MD.0000000000011803

53. Gao Y, Zheng J, Li Y, et al. Decreased functional connectivity and structural deficit in alertness network with right-sided temporal lobe epilepsy. Med (United States). 2018;97(14):1–7. doi:10.1097/MD.0000000000010134

54. Whelan CD, Altmann A, Botía JA, et al. Structural brain abnormalities in the common epilepsies assessed in a worldwide ENIGMA study. Brain. 2018;141(2):391–408. doi:10.1093/brain/awx341

55. Damoiseaux JS. Effects of aging on functional and structural brain connectivity. Neuroimage. 2017;160(January):32–40. doi:10.1016/j.neuroimage.2017.01.077

56. Dennis E, Thompson P. Functional Brain Connectivity using fMRI in Aging and Alzheimer’s Disease Emily. Neuropsychol Rev. 2014;24(1):49–62. doi:10.1007/s11065-014-9249-6. Functional

57. Hughes EJ, Bond J, Svrckova P, et al. Regional changes in thalamic shape and volume with increasing age. Neuroimage. 2012;63(3):1134–1142. doi:10.1016/j.neuroimage.2012.07.043

